# Multivariate deep phenotyping reveals behavioral correlates of non-restorative sleep in 22q11.2 Deletion Syndrome

**DOI:** 10.1101/2025.01.08.24319440

**Authors:** Natacha Reich, Andrea Imparato, Jacinthe Cataldi, Niveettha Thillainathan, Farnaz Delavari, Maude Schneider, Stephan Eliez, Francesca Siclari, Corrado Sandini

## Abstract

Converging evidence suggests that sleep disturbances can directly contribute to a transdiagnostic combination of behavior and neurocognitive difficulties characterizing most forms of psychopathology. However, it remains unclear how the growing comprehension of sleep neurophysiology should best inform sleep quality assessment in mental health patients.

To address this fundamental question, we performed deep multimodal sleep and behavioral phenotyping in 37 individuals at high genetic risk for psychopathology due to 22q11.2 Deletion Syndrome (Mean age:19±8.17, M/F=22/15) and 34 Healthy Controls (Mean age:17.06±6.87, M/F=12/22). We implemented a multivariate analysis pipeline informed by the current neurobiological understanding of the behavioral consequences of sleep disruption.

We detected multivariate patterns of disrupted sleep architecture consistently influenced by age and diagnosis across recordings and experimental settings. With high-density EEG polysomnography we detected atypical trajectories of Slow-Wave-Activity (SWA) reduction, influenced by age and sleep duration which, according to the Synaptic-Homeostasis-Hypothesis, could reflect combined alterations in neurodevelopmental and synaptic homeostasis mechanisms in 22q11DS. Blunted SWA reduction was linked with EEG markers of residual sleep pressure in morning-vs-evening EEG and with questionnaires estimating subjective somnolence in everyday life, potentially representing a clinically relevant signature of non-restorative sleep. Moreover, blunted SWA decline was linked to a transdiagnostic combination of behavioral difficulties, including negative psychotic symptoms, ADHD symptoms, and neurocognitive impairments in processing speed and inhibitory-control.

These findings suggest that systematic screening and management of sleep disturbances could directly improve behavioral outcomes in 22q11DS. They highlight the potential of precision/multivariate phenotyping approaches for characterizing the role of sleep disturbances in developmental psychopathology.

## Introduction

Sleep difficulties are highly prevalent among individuals with diverse forms of psychopathology, yet they are often overlooked or dismissed as secondary, nonspecific symptoms (1, 2). However, it is increasingly clear that sleep dysregulation can directly contribute to transdiagnostic behavioral difficulties characterizing most forms of psychopathology, including emotional dysregulation (3, 4), motivational difficulties (5), and neurocognitive attentional/executive impairments (6, 7). This is supported by a growing understanding of the neurobiological mechanisms through which sleep disturbances affect behavior (8-10). In particular, the Synaptic Homeostasis Hypothesis (SHY) proposes that slow-wave-sleep (SWS) plays an essential role in promoting brain plasticity by restoring synaptic homeostasis following learning-related synaptic strengthening occurring during wakefulness (11). According to SHY, synaptic strengthening during wakefulness results in increased neuronal synchronization that is reflected in the increased frequency and amplitude of slow-waves (SW) during sleep (11). Increased Slow-Wave-Activity (SWA) following wakefulness is therefore considered a direct marker of sleep pressure (11, 12), while SWA reduction during deep sleep could represent a proximal neurobiological measure of restorative sleep properties (11).

However, from a clinical mental health perspective, it remains largely unclear how this detailed understanding of sleep neurophysiology should best be integrated in sleep assessment and management guidelines (13). In particular, current psychiatric sleep assessments rely strongly on subjective descriptions of difficulties in initiating or maintaining sleep, which is for instance sufficient for a clinical diagnosis of insomnia (14). However, behavioral correlates of disrupted sleep can be observed in the absence of insomnia complaints (15). Indeed, complaints of insomnia can actually be anticorrelated with non-restorative sleep (16) due to the inverse relationship between sleep pressure and sleep latency (17). This underscores the need for a multimodal approach that combines subjective and objective sleep assessments with comprehensive behavioral evaluations, to accurately characterize the impact of sleep disturbances on psychopathology (18, 19). Adding to the complexity, the pathophysiology of most psychiatric disorders, including psychosis, might be traced back to neurodevelopmental periods (20, 21), during which sleep parameters undergo significant developmental changes (21, 22). For instance, SWA undergoes a dramatic 60-70% reduction during adolescence (23), mirroring a reduction in synaptic density (21, 24). Atypical adolescent synaptic pruning is highly implicated in the pathophysiology of psychosis (25, 26), which might be reflected in atypical trajectories of SWA maturation. Such multiplicity of neurodevelopmental and plasticity-related mechanisms implies that multivariate analysis techniques might be essential to characterize the associations between sleep and emergent psychopathology (13). However, performing the comprehensive sleep and behavioral phenotyping required for such multivariate analyses at early stages of psychopathology has proven challenging.

The 22q11.2 Deletion Syndrome (22q11DS) provides a unique model to explore the role of sleep disturbances in emergent psychopathology. 22q11DS is a predominantly ex-novo genetic disorder characterized by a complex multi-system somatic phenotype (27, 28), atypical neurocognitive trajectories (29, 30), and a dramatically increased risk of psychiatric disorders, including 30-40% risk of attention-deficit/hyperactivity disorder (ADHD) and anxiety disorders during childhood and a 30-40% prevalence of psychotic disorder by adulthood (31). Of note, 22q11DS is typically diagnosed at a young age, providing a unique opportunity to characterize the role of sleep disturbances in complex transdiagnostic clinical trajectories. To this date, only few studies have investigated sleep in this population, and have mostly focused on either objective or subjective sleep parameters. Subjective sleep studies highlight high prevalence of daytime somnolence, and inadequately restful sleep (32-34). Meanwhile, there is evidence that 22q11DS might be associated with high prevalence of Obstructive Sleep Apnea (OSA), due to a combination of velopharyngeal abnormalities, retrognathia and hypotonia (35-37). However, the relationship of objective and subjective sleep alterations with behavioral difficulties remains largely unclear. In a previously published study, we performed a first combined characterization of objective actigraphy-estimated sleep patterns and subjective sleep reports, which we analyzed with dedicated multivariate analysis approach to dissect behavioral correlates (34). Results suggested that 22q11DS individuals presented increased subjective somnolence despite increased sleep duration, which was suggestive of non-restorative sleep, and which further predicted psychosis vulnerability through an affective dysregulation pathway (34).

Here, we aimed to characterize the neurobiological mechanisms affecting the restorative properties of sleep in 22q11DS and their contribution to clinical and neurocognitive difficulties with the goal of informing future sleep-targeted interventions. Using a comprehensive multimodal phenotyping approach, we combined ecological and high-density polysomnography with assessments of daytime somnolence, sleep pressure, and detailed neurocognitive and psychiatric evaluations. We designed a dedicated multivariate analysis pipeline to detect developmental sleep patterns that would differentiate individuals with 22q11DS from healthy controls (HCs) and correlate with transdiagnostic behavioral difficulties. Firstly, we hypothesized that 22q11DS would show atypical sleep architecture from an early age, partly driven by OSA-related sleep fragmentation. We further hypothesized that SWA trajectories would be altered in 22q11DS, influenced by age and sleep duration, reflecting neurodevelopmental and synaptic-plasticity mechanisms. Additionally, we hypothesized that atypical sleep signatures would contribute to subjective daytime somnolence, elevated sleep pressure and transdiagnostic behavioral difficulties in motivation, emotional regulation, and neurocognitive performance. More broadly, we hypothesized that a precision multivariate phenotyping approach would be instrumental in dissecting the role of sleep dysregulation in emergent psychopathology.

## Methods

### Participants

Participants were recruited from an ongoing longitudinal study. HCs were primarily non-affected siblings (32/34). All participants and parents provided written informed consent. A total of 71 participants were included, 37 with 22q11DS (Mean age: 19±8.17, age-range: [6.72-35.54], M/F=22/15) and 34 HCs (Mean age: 17.06±6.87, age range: [6.11-37.04], M/F=12/22, see table 1).

**Table 1.** Demographic information. Differences in age were tested using a two-samples t-test and gender differences were tested using a Chi-squared test. Gender differed significantly between the two groups for the night and resting-states recordings. Age differed significantly between the two groups for the resting-states. No significant difference in any other characteristics.

|  | Night recording |  | p-value | Resting states |  | p-value |
| --- | --- | --- | --- | --- | --- | --- |
|  | 22q11DS | HC |  | 22q11DS | HC |  |
| Age: mean (SD) | 19 (8.17) | 17.06 (6.87) | 0.29 | 21.6 (6.6) | 17.53 (6.91) | **0.037** |
| Gender (M/F) | 22/15 | 12/22 | **0.041** | 15/11 | 7/18 | **0.032** |

### Clinical and Neurocognitive Assessment

22q11DS participants underwent the Structured Interview for Psychosis-Risk Syndromes (SIPS) (29 22q11DS, 20.84 ± 7.29 years old, age range [9.31 – 35.54] (38), administered by a single trained psychiatrist (SE). The SIPS yields 19 items across 4 symptom domains (positive, disorganized, negative, and general symptoms), rated on a 1-6 scale according to their severity.

Intelligent quotient (IQ) was measured using the Wechsler Intelligence Scale for Children (WISC-IV or V) (39) and Adults (WAIS-III or IV) (40). Impulsivity and inattention were measured the Connor’s Continuous Performance Test (CPT-3) (41) considering age-normed t-scores for Omission, Commission and Perseveration errors, Hit Reaction Time (HRT), HRT Inter-Stimulus Interval (ISI) Change, HRT Block change, Variability and Response Style.

Participants completed sleep questionnaires about their everyday life: the Epworth Sleepiness Scale (ESS) (42), to assess daytime somnolence as well as the Morningness and Eveningness Questionnaire (MEQ) (43), to assess circadian rhythm.

### Sleep procedure

During the first night, sleep architecture was assessed using minimally invasive Dreem Headband 2 devices(44) in an ecological setting (hotel room), followed by one-night polysomnography in a sleep laboratory designed to mimic a home environment. Parents slept in adjacent or in the same room if additional reassurance was required. Recordings started between 9 pm and 12 pm, aligning with individual sleep schedules. Before the night recordings and immediately following awakening, 5-minute eyes-closed resting-state (RS) EEGs were recorded. Both evening and night RS recordings were available for 26 participants with 22q11DS (Mean age: 21.6±6.6, age-range: [7.67-34.04], M/F=15/11) and 25 HCs (Mean age: 17.53±6.91, age-range: [6.11-37.04], M/F=7/18, see table 1).

High-density EEG was acquired with a Masti-EGI 128-channel system (https://www.magstim.com/product/long-term-eeg-sensor-nets-gel) for both resting-state and sleep recordings (sampling rate: 250 Hz). Respiratory parameters were monitored using chest and abdominal belts, a nasal cannula, and a finger oximeter, using the Masti-EGI physio16 hardware (https://www.egi.com/research-division/hardware-and-software/physiological-measurement

Sleep scoring was performed over 30 seconds epochs according to standard criteria (45). Each night was scored by two trained researchers. For the epochs where the researchers disagreed (11,72% of epochs), the scoring was determined by a validated automatic sleep scoring algorithm (https://github.com/raphaelvallat/yasa/blob/master/notebooks/14_automatic_sleep_staging.ipynb) (46). (See Supplementary table 1).

Respiratory events were evaluated by a single trained physician (CS) according to standard criteria (47), resulting in an Apnea-Hypopnea-Index (AHI) measuring the number of Apnea or Hypopnea per hour of sleep. AHI was divided by age-appropriate diagnostic cut-off for moderate OSA, corresponding to AHI>=15 (> 12 years old) and AHI >=5 (< 12 years old) (48, 49).

### Sleep and Wake Resting-State EEG analysis

First, EEG signals underwent bandpass filtering (0.5-45 Hz). Then, visually identified bad channels were excluded and interpolated using spherical splines interpolation. Independent component analysis (EEGLAB) was employed to remove potential artifacts (50).

The power spectrum of sleep recordings was computed using Fast Fourier Transform (FFT) with 2-second Hamming windows, achieving a frequency resolution of 1 Hz. From this, Delta band power (1.0–4.0 Hz) was extracted. Resting-state power spectrum analysis was performed via FFT using Welch’s sliding periodogram with a 4-second sliding window and 0.5 Hz frequency resolution, using a bandpower spectral analysis pipeline (https://github.com/raphaelvallat/yasa/blob/master/notebooks/08_bandpower.ipynb) (46). Theta relative to Alpha power was derived and used as a marker of microsleep to evaluate sleep pressure (51-55).

Additionally, slow-wave density was employed as an secondary marker of sleep pressure (56, 57). Our approach of SW detection in wake was informed from previous literature (58, 59). EEG signals were re-referenced to the average of all electrodes. SW were identified by detecting negative signal deflections between two consecutive zero-crossings, focusing on half-waves within the 1–4 Hz frequency range. To account for reduced SW amplitude during wake, we applied a relative amplitude threshold (top 10% of waves), calculated separately for each channel and participant. Delta wave density was defined as the number of waves per minute for each electrode.

### Statistical analysis

#### Comparison of Slow-Wave-Activity Trajectory during the night

We employed Mixed-Model-Analysis (MMA), implemented in MATLAB to compare SWA trajectories across the night. SWA, estimated from Power-Spectrum-Analysis, was modeled with fixed effects for night duration, age, diagnosis (22q11DS vs HC), and their interactions (age-by-duration, age-by-diagnosis, diagnosis-by-duration, and age-by-diagnosis-by-duration). Random effects included intercepts for variation in SWA across subjects and slopes for variation in SW power across subjects throughout the night, resulting in the model: (Power ∼ time + diagnosis + age + age:diagnosis + age:time + diagnosis:age + age:time:diagnosis + (1 + Time|ID)). MMA was applied independently to each of the 129 EEG channels, yielding 7 estimates per channel. Such 7×129 was then analyzed through K-Means clustering and Gap evaluation criteria, to identify groups of channels with similar variation across diagnosis, night duration, and age.

#### Detection of multivariate Sleep-Behavior correlation patterns

We employed a publicly available Partial-Least-Square-Correlation (PLSC) algorithm (https://github.com/MIPLabCH/myPLS) to identify multivariate correlations between sleep and clinical patterns. PLSC uses singular value decomposition to reveal relationships between data types, represented as bar plots where bar height and direction indicate individual variable contributions (positive vs. negative). Statistical significance of patterns was tested via permutation, while bootstrapping identified consistently contributing variables (highlighted in yellow). PLSC scores, reflecting participant-specific alignment with patterns, were calculated through matrix multiplication of observed variables by their loadings.

PLCS was firstly employed analysis sleep architecture. In this analysis, the sleep matrix included 8 variables (Sleep Onset Latency, relative proportion of 4 sleep stages, sleep duration, microarousals, sleep efficiency), while the behavioral matrix included diagnosis, age and age-by-diagnosis-interaction. Multivariate sleep scores, derived from the multiplication between participants’ sleep variables and pattern saliences, were correlated with age-normed AHI to explore respiratory contributions to altered sleep architecture.

In a second set of PLCS analyses, we modelled trajectories of sleep pressure markers in evening and morning RS recordings. Two different matrices consisting of 129 measures, a) theta/alpha power and b) SW density, were first corrected for age using linear regression, and then contrasted against a behavioral matrix consisting of diagnosis, time of day (evening vs morning), and time-by-diagnosis interaction. Through PLCS analysis, we also explored the association changes in sleep pressure (morning-evening delta SW density across channels) and SWA decline trajectories, estimated via Random-Slope-Coefficients (ID:time) from the MMA model. These coefficients reflect individual variation in SWA decline, indicating sleep pressure reduction efficiency per hour of sleep.

Finally, we explored the behavioral correlates of altered SWA reduction in 22q11DS. First, we modeled 22q11DS SWA, trough MMA, with fixed effects for age and night duration and random effects for intercept and slope per subject, resulting in the model: Power ∼ time*age + (1+time|ID). Random-Slope-Coefficients from 129 channels, reflecting individual variation in SWA decline, were correlated with three behavioral matrices 1) sleep questionnaires measuring subjective sleep quality in everyday life 2) Psychotic symptom intensity measured with the SIPS 3) Neurocognitive and attentional difficulties (WISC/WAIS and CPT variables).

## Results

**Table 2.** Sleep characteristics per group. TST: total sleep time, MA: microarousal, SE: sleep efficiency. Differences were tested using a two-samples t-test. N1 percentage and number of microarousals differed significantly between the two groups for the EEG recordings. N2 percentage differed significantly between the two groups for the dreem recording. No significant difference in any other characteristics.

|  | EEG<br>(n=71) |  |  | Dreem<br>(n=51) |  |  |
| --- | --- | --- | --- | --- | --- | --- |
|  | 22q11DS (n=37) | HC<br>(n=34) | p-value | 22q11DS<br>(n=32) | HC<br>(n=19) | p-value |
| N1 Latency | 37.23 (34.46) | 32.12 (24.35) | 0.48 | 37.78 (27.46) | 27.42 (13.72) | 0.13 |
| TST | 442.76 (81.02) | 428.57 (64.87) | 0.42 | 438.38 (85.59) | 455.39 (72.01) | 0.47 |
| %N1 | 4.27 (4.46) | 2.05 (1.94) | **0.009** | 5.9 (2.01) | 6.34 (2.34) | 0.47 |
| %N2 | 43.92 (0.99) | 46.48 (8.83) | 0.26 | 49.45 (10.75) | 42.73 (8.54) | **0.02** |
| %N3 | 17.68 (7.93) | 20.79 (7.76) | 0.1 | 20.07 (9.88) | 25.51 (10.94) | 0.07 |
| %REM | 15.95 (6.64) | 13.77 (7.86) | 0.21 | 24.58 (11.11) | 25.42 (7.29) | 0.77 |
| MA/hour | 9.81 (8.14) | 5.77 (2.81) | **0.008** | 6.72 (3.88) | 6.04 (2.85) | 0.51 |
| SE | 81.82 (11.14) | 83.09 (9.71) | 0.61 | 84.04 (7.0) | 87.16 (5.77) | 0.11 |

### Multivariate developmental sleep architecture patterns associated with 22q11DS

PLSC analysis identified two sleep-architecture patterns discriminating participants with 22q11DS from HCs. The first pattern was characterized by a negative effect of both age and 22q11DS, particularly on N3%, as well as sleep duration and sleep efficiency, combined with a positive effect of on REM%, sleep latency, and microarousal index (p=0.001, R=0.60, see figure 1.1).

**Figure 1:**
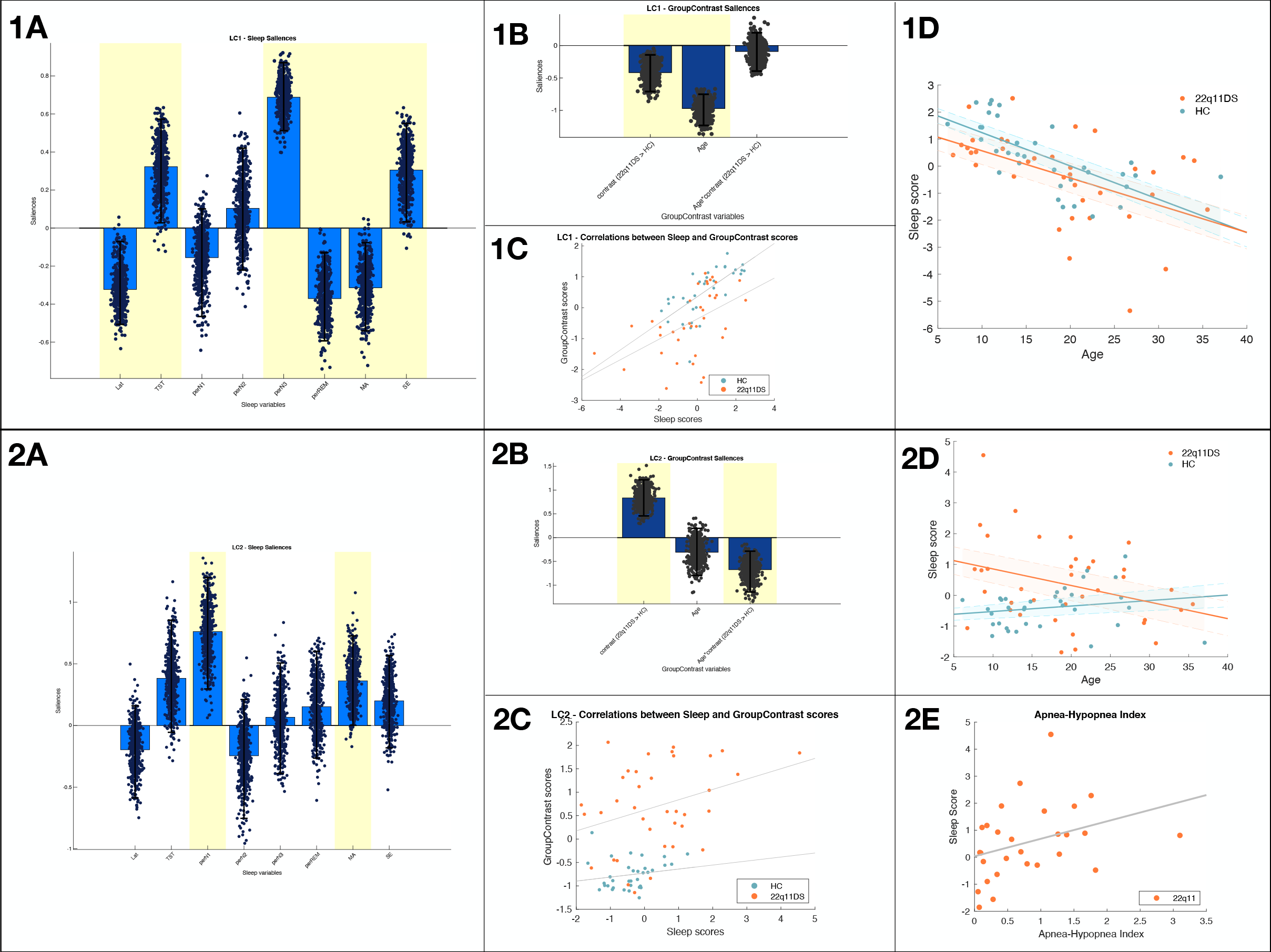
Multivariate developmental sleep architecture patterns associated with 22q11DS, two components. **Panels A:** Sleep pattern composed of sleep architecture variables. The height of each bar represents the magnitude of a variable’s contribution, while the direction (up or down) indicates whether the contribution is positive or negative. Variables consistently contributing to the pattern, as revealed by a consistent positive or negative loading within a 95% confidence interval of the bootstrapped loadings distribution, are highlighted in yellow. Scatterplots depict the distribution of a specific variable’s loadings over 500 bootstrap iterations of the original sample. **Panels B:** group contrast, age, and age-group interactions variables. As for Panels A, variables highlighted in yellow are considered to have a stable contribution to the sleep pattern, as captured by a coherent positive or negative contribution, throughout à 95% confidence interval of the bootstrapped loadings. **Panels C:** Correlation of sleep scores and group contrast scores across subjects. Each dot represents one individual. Scores reflect how each person’s variables specifically correspond to the pattern. **Panels D**: Association between individuals sleep scores and age. 22q11DS group is represented in orange whereas the HCs are in blue. **Panel 2E:** Association between sleep score and Apnea-Hypopnea Index, only 22q11DS group.

The second pattern was characterized by a positive effect of 22q11DS on N1% and microarousal index, combined with and age-by-diagnosis interaction, indicating greater sleep fragmentation in 22q11DS particularly pronounced during childhood (p=0.048, R=0.45, see figure 1.2). In 22q11DS, the score derived from this sleep-fragmentation pattern significantly correlated with the AHI, implicating respiratory events in sleep fragmentation. (see figure 1.2E, ρ=0.39, p=0.04).

### Trajectory of Slow-Wave-Activity modeled with Mixed-Models-Linear Regression

MMA revealed significant negative effects of time and age on SWA, with a positive age-by-time interaction in both groups, indicating higher SWA in younger participants and at the start of the night, with steeper declines in children. Diagnosis significantly moderated these effects, with the 22q11DS diagnosis linked to overall lower SWA and differential effects of age and time (see figure 2.1). K-means clustering of SWA trajectories highlighted two frontal and fronto-parietal clusters where the diagnosis effect was most pronounced. Temporal and occipital clusters exhibited SWA dynamics driven only by age and time, while a fifth cluster with peripheral electrodes showed no significant contributions due to low signal quality (see figure 2.2A). To further illustrate these differences, we modeled mean SWA trajectories within clusters as a function of time, divided by diagnosis and age (cutoff=18 years, see figure 2.2B). 22q11DS participants displayed blunted SWA trajectories, with relatively lower SWA at the start of the night and higher SWA at its end compared to HCs. Such atypically blunted SWA trajectory was particularly striking in younger participants, but still evident in adulthood. See supplementary materials for diagnosis, age, time, and topographic details.

**Figure 2:**
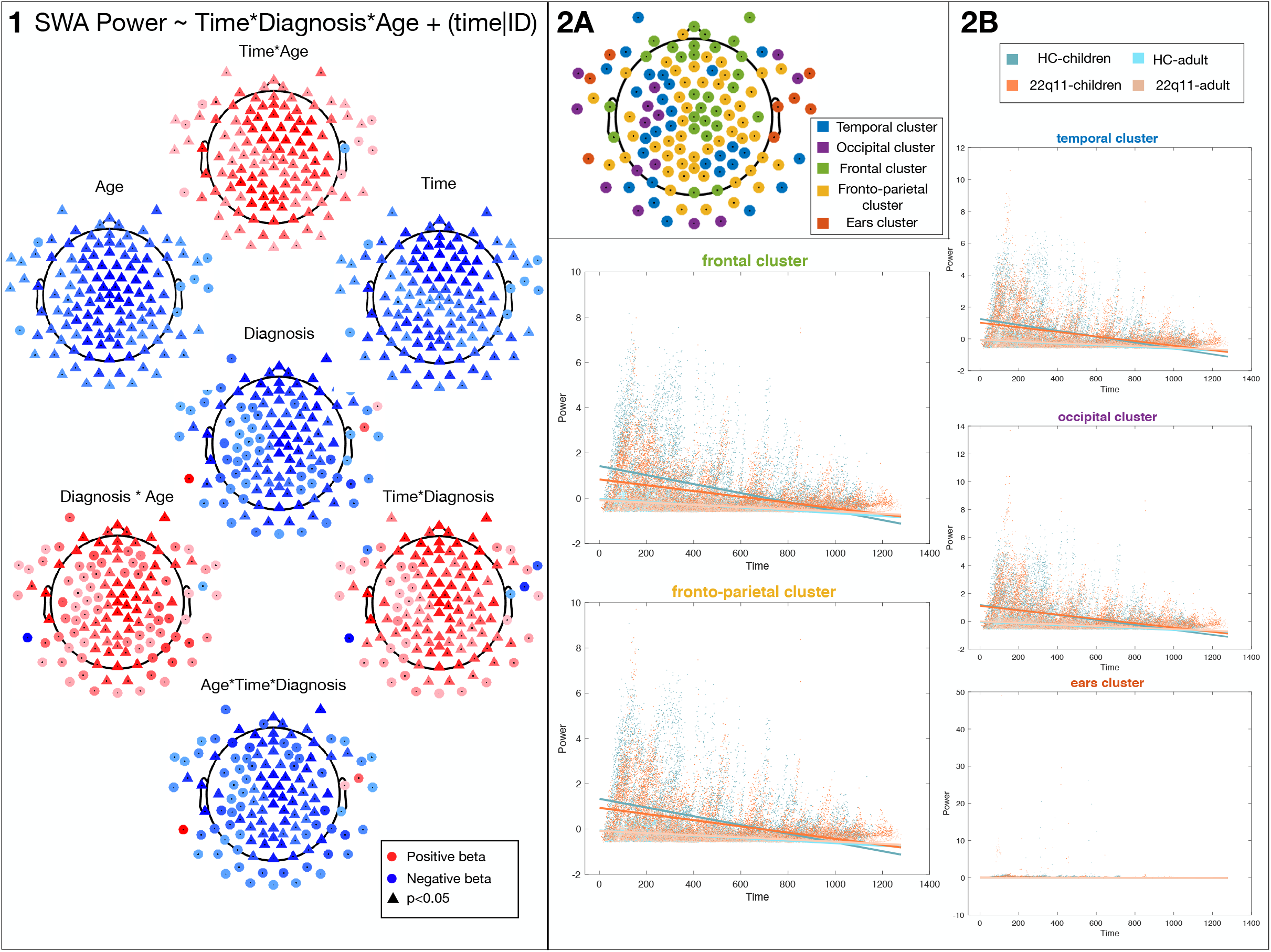
Trajectory of Slow-Wave-Activity modeled with Mixed-Models-Linear Regression. Mixed-model analysis formalized as follows: SWA Power ∼ Time*Diagnosis*Age + (time|ID) **Panel 1:** The color indicates the direction and magnitude of the estimates evaluating the contribution of each factor to SWA Power, blue represents negative estimates, and red represents positive estimates. Each electrode is marked by a shape: circles indicate non-significant contributions, while triangles denote significant contributions of the variable to the model. **Panel 2A:** Topoplot illustrating the clustering of electrodes, with each color representing a distinct cluster as identified by K-means clustering. This provides a spatial grouping of electrodes based on their similarity in model estimates. **Panel 2B:** Scatterplot illustrating the changes in power throughout the night for each of the five identified electrode clusters. Data is divided into four groups based on diagnosis and age (HC-adult/children and 22q11DS-adult/children), enabling a comparison of power dynamics across different participant groups. Scatterplots represent the z-scores of the mean power for individuals within each group, calculated for the electrodes in each cluster at each time point. The trajectories in these graphs depict how power varies across the night within each cluster and participant subgroup, derived from the estimates extracted from the mixed models of each electrode cluster.

### Trajectory of Evening vs Morning Sleep Pressure Markers in Waking Resting-State EEG

PLSC revealed that both SW-Density (p=0.005, R=0.35, see figure 3.1) and Theta/Alpha bandpower (p=0.006, R=0.28, see figure 3.2) tended to decrease in HCs, consistent with sleep pressure reduction, but presented an atypical blunted/increasing trajectory in 22q11DS, resulting in increased residual sleep pressure markers upon awakening. To investigate the link between SWA decline during sleep and residual sleep pressure, we used PLSC to contrast morning-evening SW-density changes in morning-evening Resting-State with SWA-random-slope coefficients estimated during sleep. Flatter SWA slopes correlated with reduced SW-density changes, particularly in central and fronto-parietal electrodes (p=0.006, R=0.43, see figure 3.3). This would suggest that flatted SWA slope in 22q11DS is directly linked to less efficient sleep pressure reduction, resulting in increased residual sleep pressure markers upon awakening.

**Figure 3:**
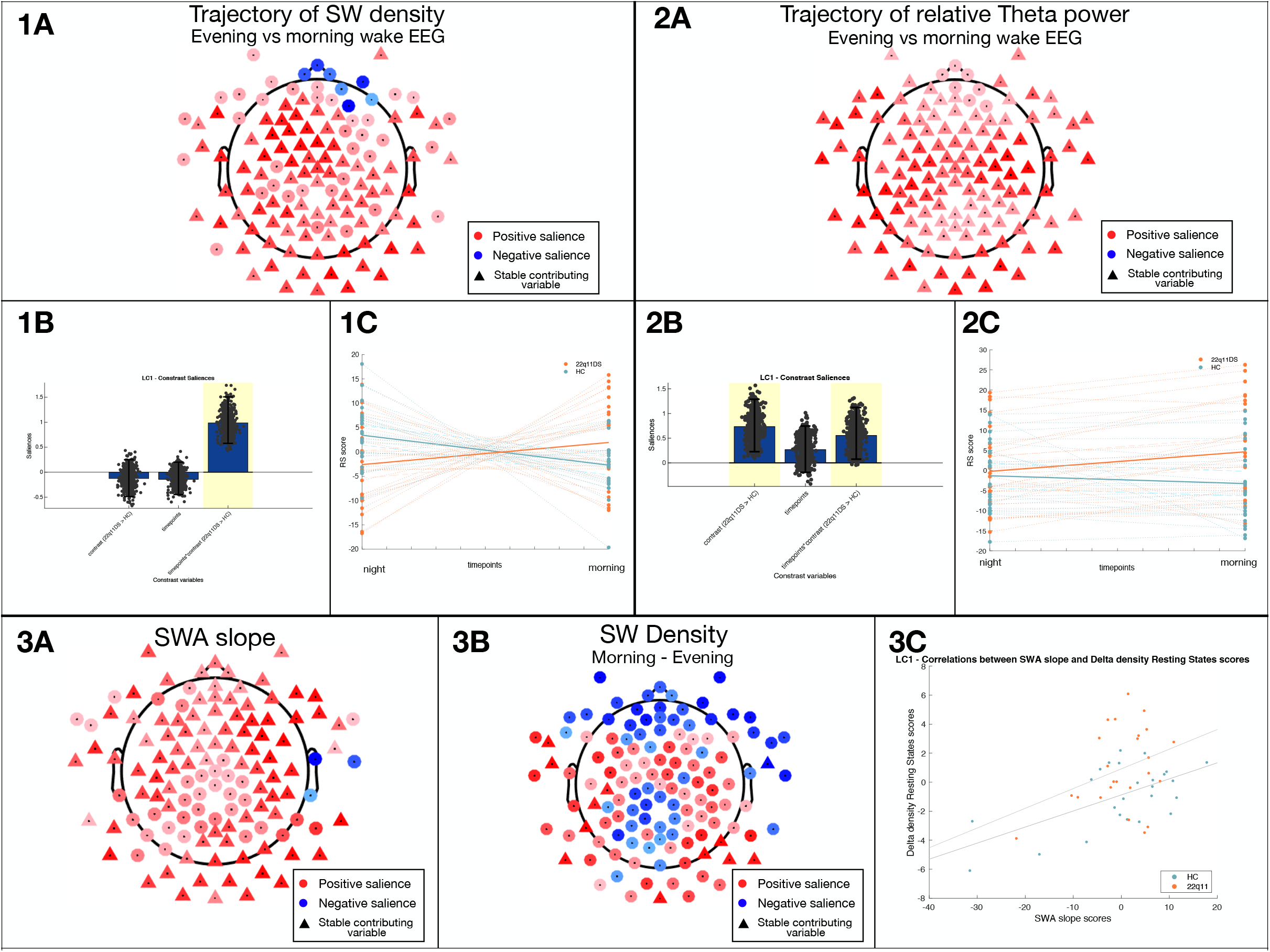
Trajectory of Evening vs Morning Sleep Pressure Markers in Waking Resting-State EEG. **Panels 1 and 2**: Multi-Variate PLSC analysis of resting states patterns associated with 22q11DS and the two timepoints. **Panel 1**: SW density at each resting state. **Panel 2**: relative theta power at each resting state. **Panels 1A:** resting states SW density. The color indicates the direction and magnitude of the loadings: blue represents negative loadings, and red represents positive loadings. Each electrode is marked by a shape: circles indicate non-stable contributions, while triangles denote stable contributions of the variable within a 95% confidence interval of the bootstrapped loadings distribution. **Panels 2A:** resting states relative theta power. The color indicates the direction and magnitude of the loadings: blue represents negative loadings, and red represents positive loadings. Each electrode is marked by a shape: circles indicate non-stable contributions, while triangles denote stable contributions of the variable within a 95% confidence interval of the bootstrapped loadings distribution. **Panels 1B and 2B:** group contrast, timepoints and timepoints-group interactions variables. Variables highlighted in yellow are considered to have a stable contribution to the sleep pattern, as captured by a coherent positive or negative contribution, throughout à 95% confidence interval of the bootstrapped loadings. Scatterplots depict the distribution of a specific variable’s loadings over 500 bootstrap iterations of the original sample. **Panels 1C and 2C**: Association between individuals sleep scores and timepoints. 22q11DS group is represented in orange whereas the HC is in blue. Panel. **Panel 3:** Multi-Variate PLSC analysis of SWA slope associated with change in resting states SW density. **Panel 3A:** Topoplot of SWA pattern composed of the random effect of the slope of each individual throughout time. The color indicates the direction and magnitude of the loadings: blue represents negative loadings, and red represents positive loadings. Each electrode is marked by a shape: circles indicate non-stable contributions, while triangles denote stable contributions of the variable within a 95% confidence interval of the bootstrapped loadings distribution. **Panel 3B:** Topoplot of change in SW density between the resting-states (morning – night). As for Panels A, variables highlighted with a triangle are considered to have a stable contribution to the sleep pattern, as captured by a coherent positive or negative contribution, throughout à 95% confidence interval of the bootstrapped loadings. **Panel 3C**: Correlation of SWA slope and delta SW density scores across subjects. Each dot represents one individual. Scores reflect how each person’s variables specifically correspond to the identified pattern. 22q11DS group is represented in orange whereas the HCs are in blue.

### Correlation of SWA Trajectory with Subjective Somnolence

To explore the association with both overall daytime somnolence as well as a potential circadian preference, we contrasted random slope coefficients of SWA decline against a behavioral matrix combining the total ESS score with individual MEQ items. Blunted SWA slopes were significantly associated with increased total ESS daytime somnolence score, combined with MEQ items capturing excessive residual morning somnolence and evening sports preference (p=0.008, R=0.48, see figure 4.1). As a confirmatory analysis, we divided participants with 22q11DS into subgroups based on their PLSC-derived EEG scores, which reflect the steepness of their SWA decline, and compared these subgroups with each other and with HCs regarding ESS and MEQ scores (60). Total MEQ scores did not significantly differ across subgroups (22q11DS flatter vs steeper trajectory: p=0.314, Cohen’s D: 0.358, 22q11DS flatter trajectory vs HCs: p=0.922, Cohen’s D: 0.0313). However, 22q11DS individuals with flatter trajectory of SWA decline presented significantly higher ESS scores both compared to 22q11DS with steeper SWA trajectory (p=0.013, Cohen’s D: -0.911), and to HCs (p=0.013, Cohen’s D: -0.794), while the latter subgroups did not significantly differ in ESS scores (p=0.544, Cohen’s D: -0.191). These results suggest that blunted SWA trajectory in 22q11DS is associated with a clinically significant excessive daytime somnolence, which manifests predominantly as increased residual morning somnolence, without being linked to an overall shift in circadian preference.

**Figure 4:**
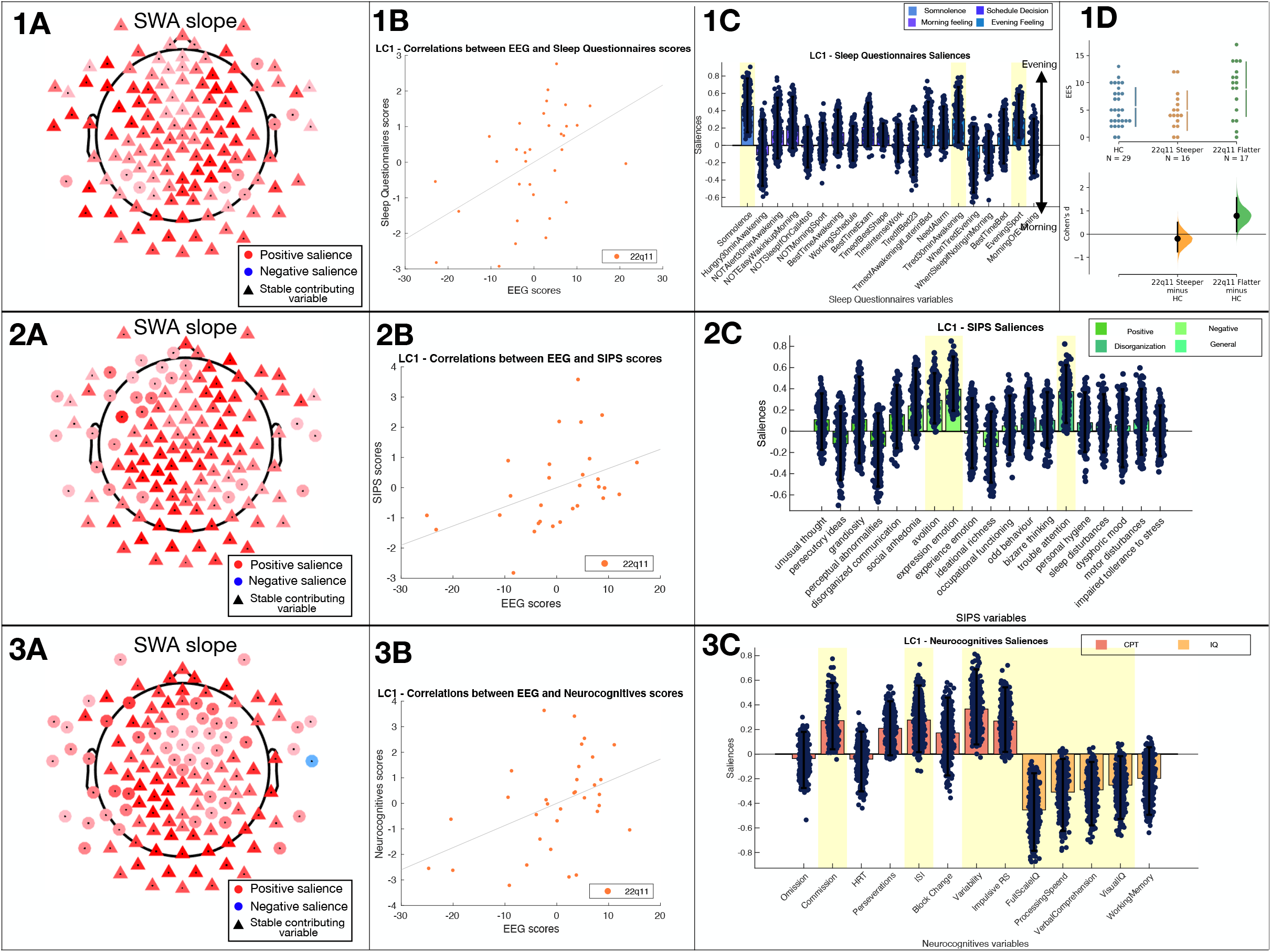
Multivariate PLSC analysis of spectral patterns associated with clinical and neurocognitive variables. **Panel 1**: Association between individual random slope and sleep questionnaires. **Panel 2**: Association between individual random slope and SIPS symptoms. **Panel 3**: Association between individual random slope and neurocognitive variables. **Panels A:** Spectral pattern composed of the random effect of the slope of each individual throughout time. The color indicates the direction and magnitude of the loadings: blue represents negative loadings, and red represents positive loadings. Each electrode is marked by a shape: circles indicate non-stable contributions, while triangles denote stable contributions of the variable within a 95% confidence interval of the bootstrapped loadings distribution. **Panels B:** Correlation of behavioral scores and EEG score scores across subjects. Each dot represents one individual. Scores reflect how each person’s variables specifically correspond to the identified pattern. **Panels C:** Clinical and neurocognitive variables. variables highlighted in yellow are considered to have a stable contribution to the sleep pattern, as captured by a coherent positive or negative contribution, throughout à 95% confidence interval of the bootstrapped loadings. Scatterplots depict the distribution of a specific variable’s loadings over 500 bootstrap iterations of the original sample. **Panel 1D**: Differences in ESS total score between HC, 22q11DS with a steeper SWA slope and 22q11DS with a flatter SWA slope. Each point represents the raw score of an individual.

### Correlation of SWA trajectories with Psychiatric and Neurocognitive Symptoms

Finally, we investigated the correlation of blunted SWA decline with psychiatric symptoms in 22q11DS, using SIPS scores. Altered SWA trajectory was associated with a clinical pattern characterized by a predominance of negative symptoms, including both avolition and reduced expressiveness, combined with more severe attentional difficulties (p=0.023, R=0.44, see figure 4.2).

Further analyses linked altered SWA decline to neurocognitive profiles, as assessed by CPT performance and WAIS/WISC IQ scores. Blunted SWA trajectories was associated with a pattern indicative of neurocognitive impulsivity, characterized by increased commission errors, higher variability, changes in ISI HRT, and a more liberal response style, which was also combined with a reduction in most of IQ subitems, particularly evident for processing speed (p=0.025, R=0.43, see figure 4.3).

## Discussion

The present study aimed to precisely characterize the sleep phenotype of 22q11DS and to assess the impact of sleep disruption on psychiatric and neurocognitive difficulties, through a unique multimodal phenotyping and multivariate analysis approach. We described atypical sleep patterns, influenced by a combination of neurodevelopmental age-related factors and sleep-dependent homeostatic mechanisms. Atypical SWA trajectories patterns were linked to excessive morning residual sleep pressure, increased daytime somnolence, and to a transdiagnostic combination of behavioral difficulties, including negative symptoms of psychosis, ADHD symptoms, and neurocognitive impairments in processing speed and inhibitory-control. These findings suggest that systematic screening and management of sleep disturbances could directly improve behavioral outcomes in 22q11DS. They highlight the potential of precision/multivariate phenotyping approaches for characterizing the contribution of sleep disturbances in developmental psychopathology (13).

A first significant observation was that sleep architecture variables varied across subjects as non-random multivariate patterns, influenced by age and diagnosis. Specifically, we detected a sleep-architecture pattern characterized by reduced sleep duration, increased sleep latency, and reduced N3%, that developed with age in both samples and was atypically increased in 22q11DS. This could suggest that neurodevelopmental mechanisms that influence age-related maturation of sleep architecture may be pathologically overrepresented in 22q11DS.

According to the SHY (61), the physiological reduction of sleep duration that disproportionally affects N3 sleep during adolescence (62, 63) is tightly linked to reduction in synaptic density and plasticity, which reduces requirements for synaptic homeostasis during SWS in adults (21). As such, atypical sleep architecture in 22q11DS could be related to reduced synaptic density and plasticity, which has been observed in animal models of the syndrome (64) and is reflected in atypical cortical maturation described by human neuroimaging studies (65). A complementary interpretation is that atypical sleep architecture in 22q11DS could reflect an inability to generate or maintain restorative SWS. Indeed, we also detected a second sleep pattern characterized by increased light sleep and in microarousals, suggesting fragmented, non-restorative sleep, which was increased in individuals with 22q11DS, particularly during childhood. This atypical sleep pattern was also correlated with the AHI, in 22q11DS, highlighting the impact of respiratory events on sleep fragmentation. Critically, these results provide a strong rational for the systematic screening of OSA since childhood in 22q11DS, which had previously gone undetected in most of our participants.

To dissect the contribution of neurodevelopmental and restorative sleep mechanisms and their relationship with behavioral difficulties, we then modelled SWA as function of both age and sleep duration. Indeed, SWA is considered as a proximal marker of synaptic-plasticity related sleep pressure (9, 12) and is also characterized by dynamic developmental trajectory that mirrors synaptic pruning during adolescence (66). Results confirmed that in both HC and 22q11DS, SWA was negatively influenced by age, in accordance with the neurodevelopmental effects of synaptic pruning (66), and by sleep duration, in accordance with the homeostatic effects of sleep (11). We also observed an interaction between the two factors, with sleep-related SWA reductions becoming less evident with increasing age, supporting the existence of more dynamic learning-related synaptic homeostasis during sleep in children compared to adults (67). Our results suggested that both age and sleep related mechanisms were atypical in 22q11DS. In particular, we observed an average reduction in SWA, particularly in fronto-parietal electrodes, in accordance with the previously described reduction in SWS. Interestingly however, SWA reductions were only observed in the early portion of the recording while SWA tended to be increased at the end of the night in 22q11DS. This was driven by a blunted trajectory of SWA reduction during the night, which was particularly striking in children. According to the SHY, such blunted trajectories of SWA reduction could indicate a reduced efficiency in achieving restorative synaptic homeostasis during sleep (9).

To confirm the behavioral relevance of SWA trajectories, we first investigated the change of sleep pressure signatures in morning compared to evening resting-state recordings. We observed an insufficient reduction in sleep pressure markers in morning compared to evening recordings in 22q11DS, evident in terms of both relative theta/alpha power and the density of SW. Furthermore, we aimed to investigate whether residual sleep pressure was directly associated with SWA across the night. Our findings indicate that a flattened SWA slope is closely linked to markers of residual sleep pressure. These results would confirm that blunted trajectory of SWA decline is linked to an insufficiently restorative reduction of sleep pressure, resulting in excessive residual sleep pressure upon awakening. To explore the clinical relevance of these findings beyond the experimental setting, we then associated SW trajectory with validated measures of daytime somnolence and circadian disturbances through a dedicated multivariate analysis. Results confirmed that 22q11DS individuals with blunted SWA trajectories presented significant increase in ESS, both compared to 22q11DS individuals with steeper SWA trajectories and to HCs. Symptoms of excessive somnolence were reported predominantly shortly after awakening, which was not accounted for by overall differences in circadian preference measures. A possible interpretation is that the subjective experience of excessive residual sleep pressure is strongest close to awakening, when the arousal system is typically less active (68). We previously reported that individuals with 22q11DS present excessive subjective somnolence despite objectively increased sleep duration, estimated through actigraphy (34). We interpreted this as evidence of non-restorative sleep, although actigraphy-based sleep phenotyping was insufficient to confirm this hypothesis (34). Here we show that increased subjective somnolence in 22q11DS is specifically related to blunted trajectories of SWA reduction.

Finally, we explored the clinical correlates of altered SWA reduction. Indeed, previous evidence suggested that subjectively non-restorative sleep longitudinally predicted worst mental health trajectories in 22q11DS (34). A multivariate analysis revealed that flattened SWA trajectories were associated with transdiagnostic behavioral difficulties, affecting negative symptoms of psychosis, including reduced motivation and blunted affect, and symptoms of attentional difficulties. Blunted SWA reduction was also associated with a pattern of neurocognitive impairment predominantly characterized by reduced processing speed and impaired top-down inhibitory control. This suggests that the consequences of less efficient sleep pressure reduction extend beyond subjective somnolence and contribute to the psychiatric and neurocognitive difficulties observed in 22q11DS. Of note, negative symptoms of psychosis are highly prevalent in 22q11DS (69) and, similarly to what reported idiopathic schizophrenia, are correlated with neurocognitive difficulties, including processing speed and inhibitory control (29, 70). The pathophysiology of such negative and neurocognitive symptoms is partially understood and likely multifactorial (71, 72). Interestingly, converging evidence suggests that insufficiently restorative sleep can influence a remarkably similar combination of behavioral difficulties, affecting motivation (5), processing speed (73, 74) and ADHD symptoms (75), due to a direct effect of sleep synaptic homeostasis mechanisms on brain networks implicated in goal directed executive functioning (5, 76) and top-down inhibitory control (59). As such, our results suggest that improving the restorative properties of SWS could directly improve a combination of negative psychotic symptoms and neurocognitive difficulties, that disproportionally influence functional impairment in 22q11DS (70) and idiopathic schizophrenia (77), and for which effective treatment options are currently lacking (78).

Overall, our results highlight the clinical potential of precision/multivariate phenotyping approaches for characterizing the mechanisms linking sleep disturbances and psychopathology. Indeed, our results suggest that sleep and psychopathology could be linked by a multivariate combination of neurodevelopmental and synaptic plasticity related mechanisms that would go undetected if individual sleep and behavioral variables were analyzed in isolation. From a clinical perspective, this provides a strong rational for improving sleep quality assessments, particularly in individuals affected by transdiagnostic psychopathology. Still, this study should be considered in light of significant limitations. Firstly, our cross-sectional design could not conclusively ascertain causal directionality between sleep and behavioral difficulties. However, we previously showed that in 22q11DS excessive somnolence predicted worsening behavioral difficulties, including in particular negative symptoms, over a 3-year longitudinal follow-up (34), while the current results strongly link atypical SWA trajectory with both residual sleep pressure upon awakening and subjective somnolence in everyday life. Secondly, we have limited understanding of the pathophysiology of altered SWA in 22q11DS, and its general relevance for non-syndromic mental health patients. For instance, velopharyngeal abnormalities associated with 22q11DS might influence respiratory events that significantly contributed to sleep fragmentation in our data (35, 37). Future studies should further investigate the contribution of respiratory events to restorative sleep properties in 22q11DS, which as detailed in supplementary material, was challenging due to our study’s wide age-range. Still, our results provide a strong rational for including systematic screening of OSA among future 22q11DS treatment guidelines (79). Indeed, growing evidence suggests that OSA might be underdiagnosed among non-syndrome mental health patients (80), and represent an effective treatment target (81). Overall, our results support future efforts at clarifying the emergence and downstream consequences of non-restorative sleep in mental health patients, by informing sleep phenotyping and analysis approaches with our current understanding of sleep physiology (13).

## Data Availability

All data produced in the present study are available upon reasonable request to the authors

