## Supplementary material for "Multivariate deep phenotyping reveals behavioral correlates of non-restorative sleep in 22q11.2 Deletion Syndrome"

**Supplementary tables:**

Disagreement between sleep scorers.

Disagreements in staging across scorer were calculated for each stage. Subsequently, the total of epochs with disagreement for each night was divided by the total number of epochs of the night (see supplementary table 1).

|  | **22q11DS** | **HC** | p-value |
| --- | --- | --- | --- |
| Ratio of epochs (mean ± std) | 0.13 ± 0.06 | 0.11 ± 0.03 | 0.07 |

*Supplementary table 1: Ratio of disagreements in staging across scorers (epochs with differences/epochs total). Statistical difference evaluated with a two-sample t-test.*

Mixed-model statistics of Slow-Wave-Activity Trajectory during the night by cluster.

We modelled the trajectory of SWA during the night for each cluster of electrodes, using a mixed-model analysis, resulting in the following model: (Power ~ time + diagnosis + age + age:diagnosis + age:time + diagnosis:age + age:time:diagnosis + (1 + Time|ID)) (see in methods for more details). We extracted the mixed-model statistics for the 5 clusters (see supplementary table 2). For each cluster, the variables contributing significantly to the model are highlighted in bold.

|  | | *Intercept* | *Time* | *Diagnosis* | *Age* | *T:D* | *T:A* | *D:A* | *T:D:A* |
| --- | --- | --- | --- | --- | --- | --- | --- | --- | --- |
| *Temporal Cluster* | *Estimate* | **2.639** | **-0.003** | -0.735 | **-0.116** | **1.1*10^-4^** | **9.89*10^-4^** | 0.037 | -4.45*10^-5^ |
|  | *SE* | **0.302** | **0.0003** | 0.408 | **0.165** | **0.0004** | **1.818*10^-5^** | 0.211 | 2.332*10^-5^ |
|  | *tStat* | **8.719** | **-9.652** | -1.8 | **-7.05** | **2.205** | **6.274** | 1.751 | -1.907 |
|  | *DF* | **48259** | **48259** | 48259 | **48259** | **48259** | **48259** | 48259 | 48259 |
|  | *pVal* | **2.902*10^-18^** | **5.05*10^-22^** | 0.072 | **1.828*10^-12^** | **0.027** | **3.54*10^-10^** | 0.08 | 0.057 |
|  | *Lower* | **2.046** | **-0.004** | -1.535 | **-0.149** | **0.0001** | **7.843*10^-5^** | -0.004 | -9.016*10^-5^ |
|  | *Upper* | **3.232** | **-0.003** | 0.065 | **-0.084** | **0.002** | **0.0001** | 0.079 | 1.242*10^-6^ |
| *Ears Cluster* | *Estimate* | 0.036 | -3.337*10^-5^ | 0.499 | -0.002 | -6.025*10^-5^ | 1.185*10^-6^ | -0.0009 | 1.248*10^-6^ |
|  | *SE* | 0.066 | 7.905*10^-5^ | 0.089 | 0.004 | 0.0001 | 4.59*10^-6^ | 0.005 | 5.715*10^-6^ |
|  | *tStat* | 0.547 | -0.422 | 0.563 | -0.562 | -0.578 | 0.258 | -0.19 | 0.218 |
|  | *DF* | 48259 | 48259 | 48259 | 48259 | 48259 | 48259 | 48259 | 48259 |
|  | *pVal* | 0.585 | 0.673 | 0.573 | 0.574 | 0.563 | 0.796 | 0.849 | 0.827 |
|  | *Lower* | -0.093 | -0.0002 | -0.124 | -0.009 | -0.0003 | -7.812*10^-6^ | -0.001 | -9.954*10^-6^ |
|  | *Upper* | 0.165 | 0.00012 | 0.223 | 0.005 | 0.0001 | 1.082*10^-5^ | 0.008 | 1.245*10^-5^ |
| *Fronto-parietal Cluster* | *Estimate* | **2.807** | **-0.003** | **-1.028** | **-0.123** | **0.001** | **0.0001** | **0.048** | **-5.429*10^-5^** |
|  | *SE* | **0.285** | **0.0003** | **0.384** | **0.016** | **0.0004** | **1.727*10^-5^** | **0.02** | **-2.215*10^-5^** |
|  | *tStat* | **9.851** | **-10.645** | **-2.674** | **-7.923** | **2.956** | **6.952** | **2.398** | **-2.452** |
|  | *DF* | **48259** | **48259** | **48259** | **48259** | **48259** | **48259** | **48259** | **48259** |
|  | *pVal* | **7.108*10^-23^** | **1.965*10^-26^** | **0.008** | **2.374*10^-15^** | **0.003** | **3.636*10^-12^** | **0.016** | **0.014** |
|  | *Lower* | **2.249** | **-0.004** | **-1.781** | **-0.153** | **0.0004** | **8.622*10^-4^** | **0.009** | **-1.09*10^-5^** |
|  | *Upper* | **3.365** | **-0.003** | **-0.274** | **-0.093** | **0.002** | **0.0002** | **0.087** | **-1.089*10^-5^** |
| *Occipital Cluster* | *Estimate* | **2.439** | **-0.003** | -0.42 | **-0.106** | 0.0007 | **0.0001** | 0.024 | -3.16*10^-5^ |
|  | *SE* | **0.305** | **0.0003** | 0.412 | **0.017** | 0.0005 | **1.89*10^-5^** | 0.021 | 2.42*10^-5^ |
|  | *tStat* | **7.99** | **-8.799** | -1.02 | **-6.389** | 1.422 | **5.571** | 1.1483 | -1.304 |
|  | *DF* | **48259** | **48259** | 48259 | **48259** | 48259 | **48259** | 48259 | 48259 |
|  | *pVal* | **1.397*10^-15^** | **1.424*10^-18^** | 0.307 | **1.682*10^-10^** | 0.155 | **2.54*10^-8^** | 0.251 | 0.192 |
|  | *Lower* | **1.84** | **-0.004** | -1.227 | **-0.124** | -0.0003 | **6.824*10^-5^** | -0.017 | -7.909*10^-5^ |
|  | *Upper* | **3.3037** | **-0.002** | 0.387 | **-0.0737** | 0.002 | **0.0001** | 0.066 | 1.59*10^-5^ |
| *Frontal Cluster* | *Estimate* | **2.951** | **-0.0003** | **-1.359** | **-0.128** | **0.001** | **0.0001** | **0.006** | **-6.211*10^-5^** |
|  | *SE* | **0.271** | **0.0003** | **0.366** | **0.015** | **0.0004** | **1.66*10^-5^** | **0.019** | **2.129*10^-5^** |
|  | *tStat* | **10.871** | **-11.262** | **-3.711** | **-8.639** | **3.654** | **7.209** | **3.158** | **-2.917** |
|  | *DF* | **48259** | **48259** | **48259** | **48259** | **48259** | **48259** | **48259** | **48259** |
|  | *pVal* | **1.708*10^-27^** | **2.188*10^-29^** | **0.0002** | **5.864*10^-18^** | **0.0003** | **5.723*10^-13^** | **0.001** | **0.004** |
|  | *Lower* | **2.419** | **-0.004** | **-2.08** | **-0.157** | **0.0007** | **8.715*10^-5^** | **0.023** | **-0.0001** |
|  | *Upper* | **3.483** | **-0.003** | **-0.641** | **-0.099** | **0.002** | **0.0002** | **0.097** | **-2.038*10^-5^** |

*Supplementary table 2: Mixed-models’ statistics. In bold, variables participating significantly to the model. T: Time, D: Diagnosis, A: Age*

**Supplementary analyses:**

Supplementary analysis 1: Habituation night: dreem headband

**Methods**

As a first supplementary analysis, we used the partial-least-square-correlation (PLSC) (see in methods for more details), to investigate possible difference between the night in an ecological setting and the night in a laboratory setting. To this end, we performed a PLSC on the hotel recordings (dreem headband) and compared the sleep architecture between group and with age. Subsequently, we correlated the sleep score extracted from the hotel night and the sleep pattern extracted from the sleep laboratory night. Only participants with the two nights were kept (19 22q11DS, mean age: 18.67 ± 7.61, age range: [6.73 -34.04] and 17 HCs, mean age: 15.56 ± 5.48, age range: [9.82-25.94]).

**Results**

Individuals with the 22q11DS differentiated from HCs by a reduction in N3%, increase in N2% and more microarousals. The individual sleep scores extracted from the dreem pattern correlated significantly with the sleep scores extracted from the EEG sleep pattern (R = 0.51, p = 0.002).

**Discussion**

There was a significant correlation between the sleep scores extracted from the PLSC conducted on the hotel night and the laboratory night. This correlation indicates that the sleep disturbances observed in the 22q11DS group are not merely artifacts of the laboratory environment but are reflective of their typical sleep architecture in a more naturalistic setting. These findings underscore the robustness and ecological validity of our results, highlighting the persistent nature of sleep disruptions in individuals with 22q11DS across different sleep environments.

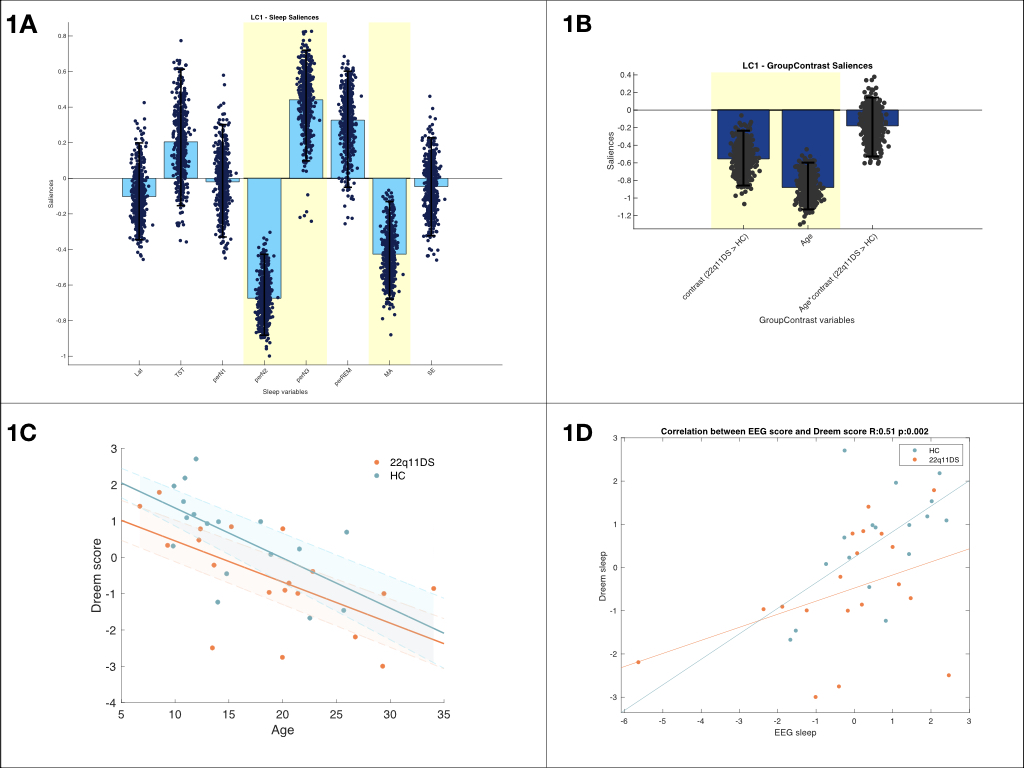

*Supplementary figure 1: Multi-Variate PLSC analysis of sleep patterns of the dreem headband.* ***Panel 1****: Multi-Variate PLSC analysis of sleep patterns associated with the 22q11DS.* ***Panel 1A:*** *Sleep pattern composed of sleep architecture variables from the dreem headband. The height of each bar represents the magnitude of a variable's contribution, while the direction (up or down) indicates whether the contribution is positive or negative. Variables consistently contributing to the pattern, as indicated by a consistent positive or negative loading within a 95% confidence interval of the bootstrapped loadings distribution, are highlighted in yellow. Scatterplots depict the distribution of a specific variable's loadings over 500 bootstrap iterations of the original sample.* ***Panel 1B:*** *group contrast, age and age-group interactions variables. As for Panels A, variables highlighted in yellow are considered to have a stable contribution to the sleep pattern, as captured by a coherent positive or negative contribution, throughout à 95% confidence interval of the bootstrapped loadings.* ***Panel 1C:*** *Association between individuals sleep scores and age. 22q11DS group is represent in orange whereas the HCs are in blue*. ***Panel 1D****: Association between individuals sleep scores extracted from the group-contrast PLS with the EEG (see figure 1.1) and group-contrast PLS with the dreem (see figure supplementary 1A-B) on the y-axis. 22q11DS group is represent in orange whereas the HCs are in blue.*

Supplementary analysis 2: Impact of gender on SWA

**Methods**

To assess the impact of gender differences on SW power, we constructed a mixed-model analysis extending the primary model detailed in the main text by incorporating gender and the interaction term gender:diagnosis as fixed effects. The resulting model was Power ~ Time*Diagnosis*Age + Gender*Diagnosis + (1 + Time|ID). Consistent with the main text, this mixed-model-analysis was applied independently to each channel, yielding 9 estimate values for 129 channels. These estimates were then visualized using 9 topoplots to provide spatial insights.

**Results**

The analysis revealed that neither gender nor the interaction between gender and diagnosis contributed significantly to the model variance (see supplementary figure 2).

**Discussion**

Our findings indicate that gender differences did not significantly influence SW power, nor did the interaction between gender and diagnosis. Therefore, gender was not included as a variable in the primary analyses. This supports the robustness of our results, as gender does not appear to confound the observed differences SWA between the 22q11DS and HC groups.

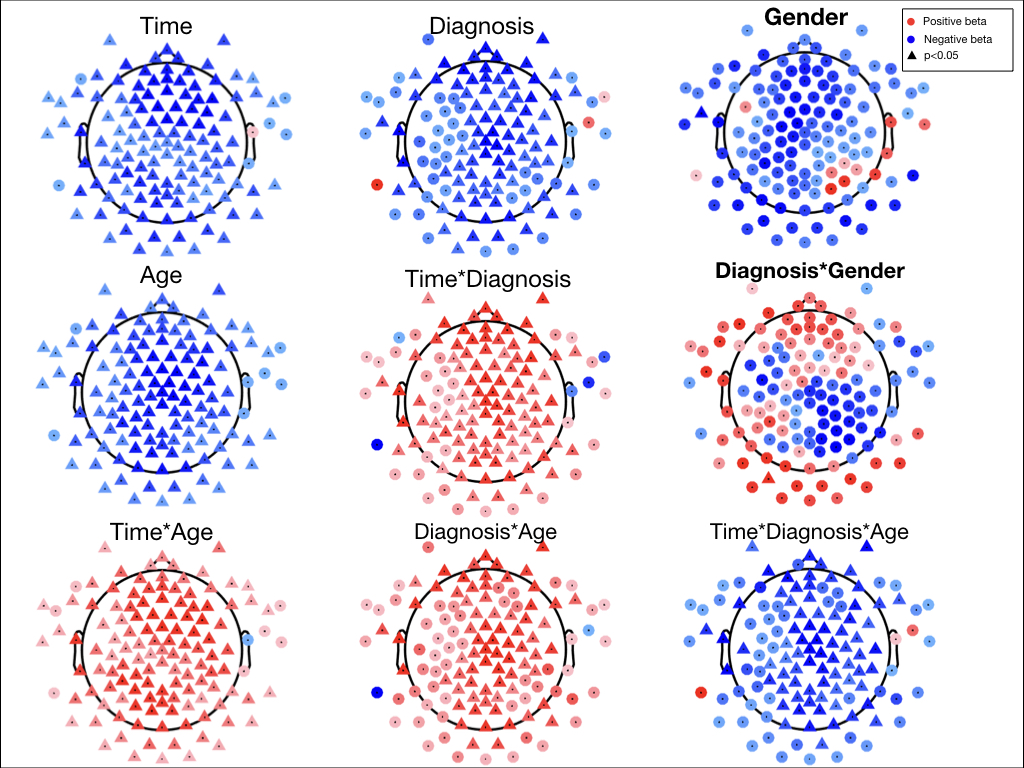

*Supplementary figure 2: Mixed-model analysis Power ~ Time*Diagnosis*Age + Diagnosis*Gender + (time|ID):* *The colour indicates the direction and magnitude of the estimates: blue represents negative estimates, and red represents positive estimates. Each electrode is marked by a shape: circles indicate non-significant contributions, while triangles denote significant contributions of the variable to the model. Topoplots associated with gender are highlighted in bold.*

Supplementary analysis 3: Association between Apnea-hypopnea episodes and clinical PLS

**Methods**

As a supplementary analysis, we examined the relationship between patterns identified by the clinical PLS analysis (see figure 4) and apnea-hypopnea episodes, reflected by the age-normed Apnea-Hypopnea Index (AHI). The individual scores reflect the degree to which a participant's variables correspond to the EEG pattern (see Figure 4A) or the clinical pattern (see Figure 4C) identified by the PLS. To account for AHI, we first adjusted both EEG and clinical scores for age-normalized AHI using linear regression and subsequently assessed the association between the adjusted scores using Pearson correlation.

**Results**

The correlations between the EEG and clinical scores remained significant after adjusting for AHI (R=0.56, p=0.003; R=0.50, p=0.015; and R=0.49, p=0.012; see Supplementary Figure 3A). Age normed AHI scores were not significantly correlated with either multivariate EEG scores estimating SWA slope trajectory over the night.

**Discussion**

These results would suggest that blunted SWA trajectory decline and its correlation with behavioural outcomes is not entirely accounted for by the frequency of respiratory events associated with Obstructive Sleep Apnea. This could suggest a multifactorial origin of atypical SWA trajectories in 22q11DS. It is however important to note that several methodological limitations hinder the ability to draw strong conclusions on the contribution of respiratory events. Firstly, a significant proportion participants (9/37), did not tolerate wearing nasal cannulas required for estimation of OSA. Secondly the AHI clinical thresholds estimating severity of OSA change dramatically from paediatric to adult populations [1]. In order to establish the impact of OSA on SWA trajectories it would therefore be important to conduct separate analysis in children and adults with 22q11DS, specifically investigating how the correlation between AHI and SWA evolves with age, which our study is underpowered to do. Future studies should specifically investigate the impact of respiratory events on SWA trajectories for instance by investigating whether OSA treatment could have beneficial effects of objective and subjective signatures of non-restorative sleep in 22q11DS, employing a within-subject longitudinal design.
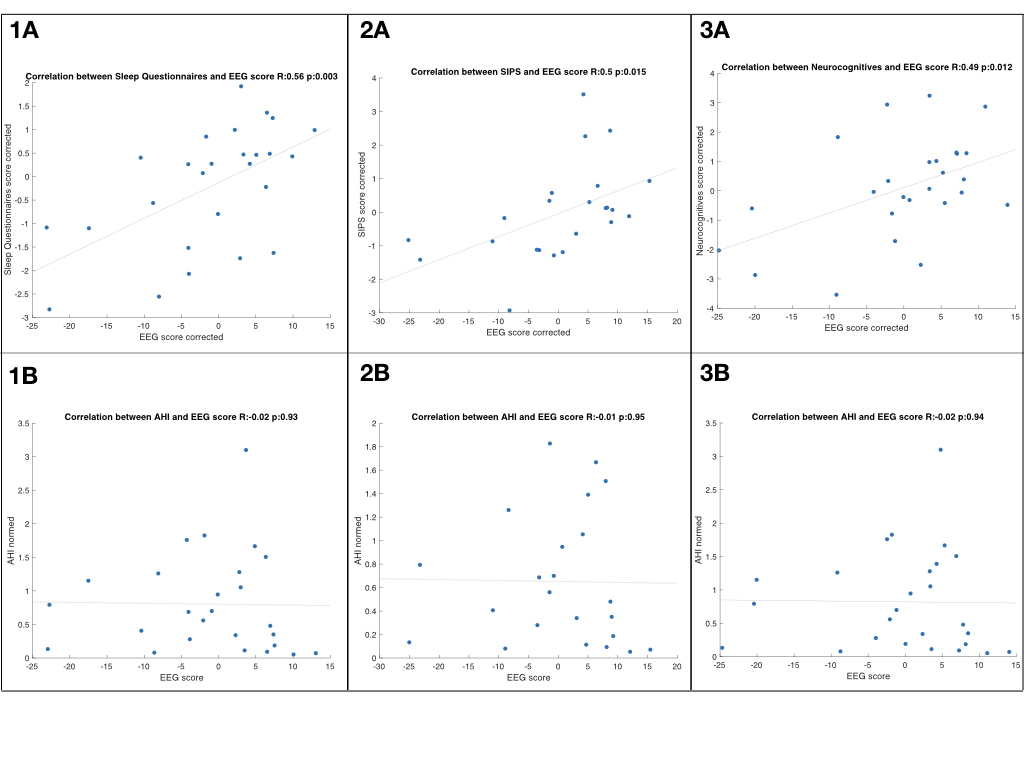

*Supplementary figure 3: Association between the clinical PLS patterns and the AHI****. Panel 1:*** *Sleep questionnaires, individual EEG spectral slope and AHI.* ***Panel 2****: SIPS symptoms, individual EEG spectral slope and AHI.* ***Panel 3:*** *Neurocognitive variables, individual EEG spectral slope and AHI.* ***Panels A****: Correlation between clinical scores and EEG scores, after adjusting for AHI.* ***Panels B****: Correlation between AHI and EEG scores.*

Supplementary analysis 4: Slow wave activity in resting states

**Methods**

Here, we report on results of SW detection as well as relative theta power analysis in evening vs morning waking resting-state recordings. In addition to SW-density values, which we considered as a direct marker of sleep pressure and analysed in the main text, we report results of SW peak-to-peak amplitude. Moreover, in addition to multivariate PLSC described in the main text we performed univariate comparisons of SW-density, PTP amplitude and relative theta power in evening and morning RS recordings as well as change evening-minus-morning values across HC and 22q11DS populations using two-sample t-tests. A graphical representation of these results is provided in Supplementary Figures 4 and 5.

**Results**

In the evening, there was a lower density of SW in the 22q11DS compared to the HCs in most of the electrodes and higher mean PTP amplitude in most of the frontal and occipital electrodes (see supplementary figures 4A). There were no significant differences in relative theta power across groups in the evening (see supplementary figures 5A). In the morning, individuals with 22q11DS exhibited a significantly higher mean slow wave density in several frontal electrodes compared to HC, higher mean PTP in a few frontal electrodes (see supplementary figures 4B) as well as higher relative theta power in most of the electrodes (see supplementary figures 5B). When analysing the change from night to morning (delta), individuals with 22q11DS showed positive delta in terms of SW density (see supplementary figure 4.3.C) and relative theta power (see supplementary figure 5.1.C), whereas HC showed a negative delta (see supplementary figures 4.4.C and 5.2.C), with most electrodes significantly different between groups. No significant differences were observed between groups in SW amplitude delta across electrodes (see supplementary figures 4.1.C and 4.2.C).

**Discussion**

These univariate results are in line with results multivariate PLCS analysis described in the main text and highlight differential trajectory of SW in 22q11DS. The elevated slow wave density and relative theta power observed in the 22q11DS group in the morning, supports the notion that they may not be as well-rested, with sleep pressure remaining higher even after a night of sleep. Additionally, the lack of significant differences between morning and evening resting-states (delta) in SW amplitude suggests that the higher sleep pressure in 22q11DS may be reflected more in the density of slow waves than in their intensity. These results provide evidence that individuals with 22q11DS may have difficulty achieving restorative sleep, leading to increased sleep pressure the following morning.

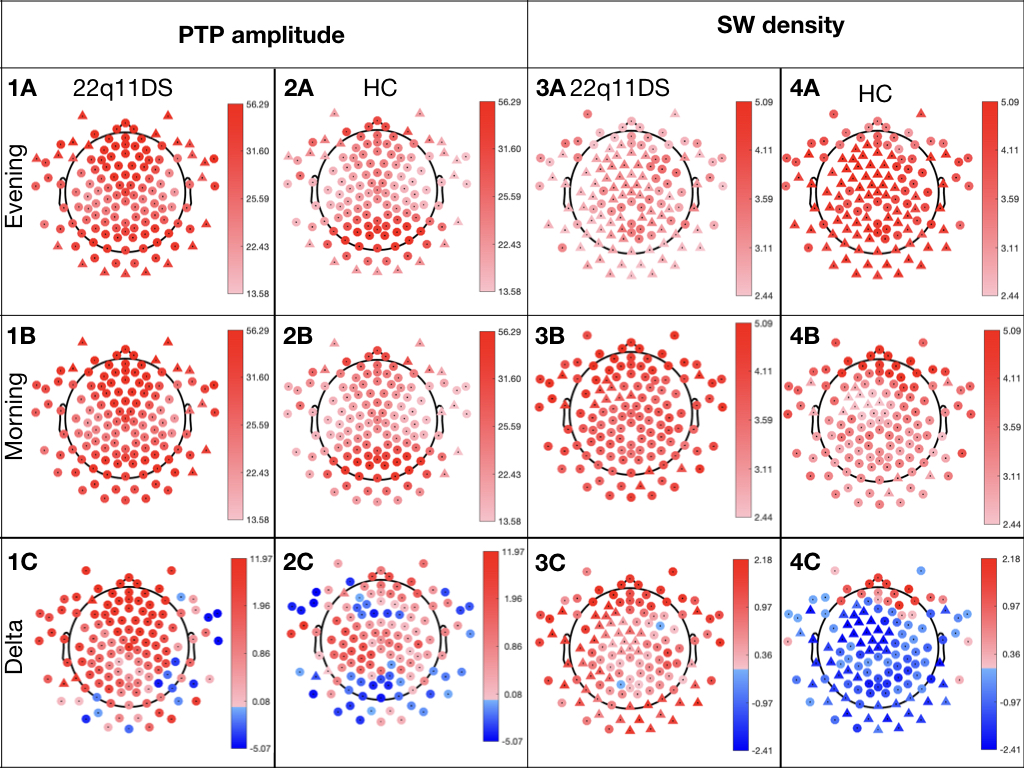

*Supplementary figure 4: Slow-waves detection on morning and evening resting-states. Differences between groups were tested using two-samples t-test. Electrodes with a significant difference are represented with a triangle. For each variable, the mean was calculated by electrode and by group.* ***Panels 1:*** *mean 22q11DS Peak-to-Peak amplitude (μV).* ***Panels 2:*** *mean HC Peak-to-Peak amplitude (μV).* ***Panels 3:*** *mean 22q11DS Slow-wave density (SW/min).* ***Panels 4:*** *mean HC Slow-wave density (SW/min).* ***Panels A:*** *Evening resting-state.* ***Panels B:*** *Morning resting-state. Panels C: Delta of mean values of RS (morning – night).*
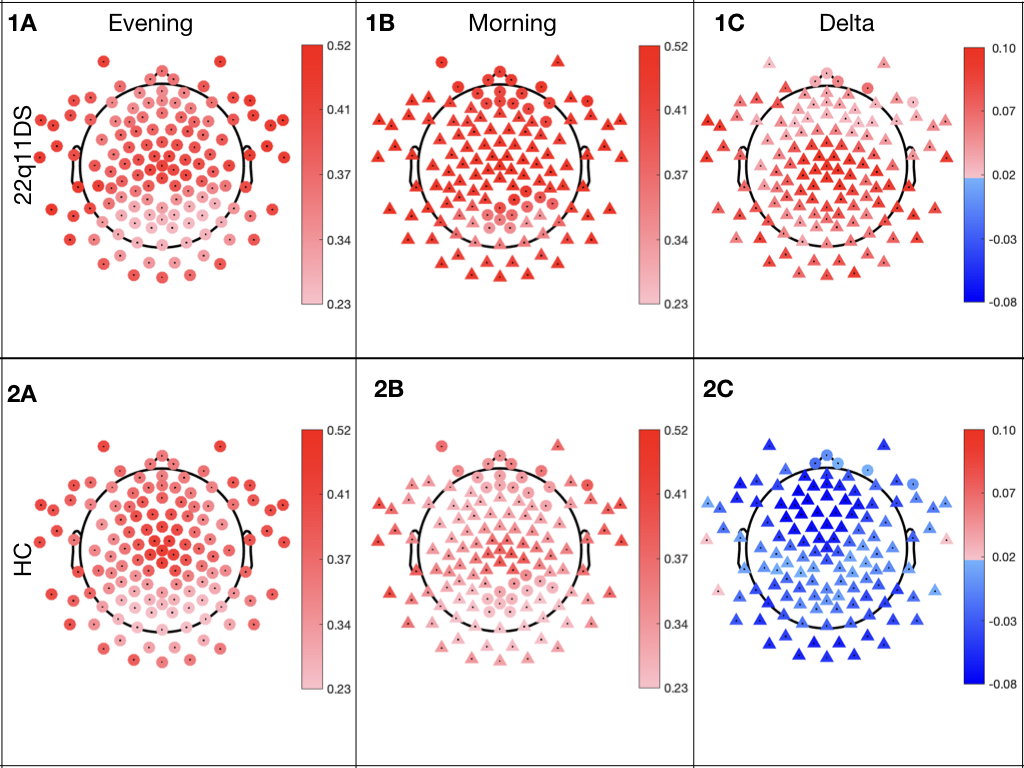

*Supplementary figure 5: Relative theta power in morning and night resting-states. Differences between groups were tested using two-samples t-test. Electrodes with a significant difference are represented with a triangle, red indicates positives value and blue negative values. For each variable, the mean was calculated by electrode and by group.* ***Panel 1:*** *22q11DS.* ***Panel 2:*** *HC.* ***Panels A****: Evening resting-state.* ***Panels B:*** *Morning resting-state.* ***Panels C:*** *Delta of mean values of RS (morning – night).*

Supplementary analysis 5: Slow wave power

**Methods**

For descriptive purposes, we represented SWA as function of age across diagnostic groups. SWA is averaged across electrodes clustered according to differential SWA trajectories across groups, yielding 5 topographical clusters: Temporal, Peripheral/Peri-Auricular, Frontal, Fronto-Parietal and Occipital. See Methods; Statistical Analysis: Comparison of Slow-Wave Activity Trajectory during the Night. Within such clusters, SW power was averaged across the entire night, as well as in the first and last 30 minutes of NREM sleep (both N2 and N3 stages). Additionally, we computed the change in power (delta) between start and end of night. Quantitative results of this analysis are reported in Supplementary Table 3 for both HC and individuals with 22q11DS, each split by age (children < 18 and adults). Moreover, average SW power results are plotted as a function of age in Supplementary Figure 6.

**Results**

In accordance with results reported in the main text SW-Power was strongly negatively influenced by age in both HCs and 22q11DS individuals. Groups differences were most striking for change in SW-Power across the night, with stronger SW-Power reduction in HCs compared to 22q11DS, particularly during childhood, and particularly in frontal and fronto-parietal clusters (Panels 4D and 5D). This differential trajectory resulting in higher end-night SW-Power in 22q11DS (Panels 4C and 4D) as opposed to a reduction observed at the start of the night (Panels 3C and 3D).

**Discussion**

These results highlight a flatter reduction in SWA particularly in young individuals with 22q11DS, in agreement with results of the mixed-model analysis described in the main text, and potentially indicative of inefficient reduction of sleep pressure during the night.

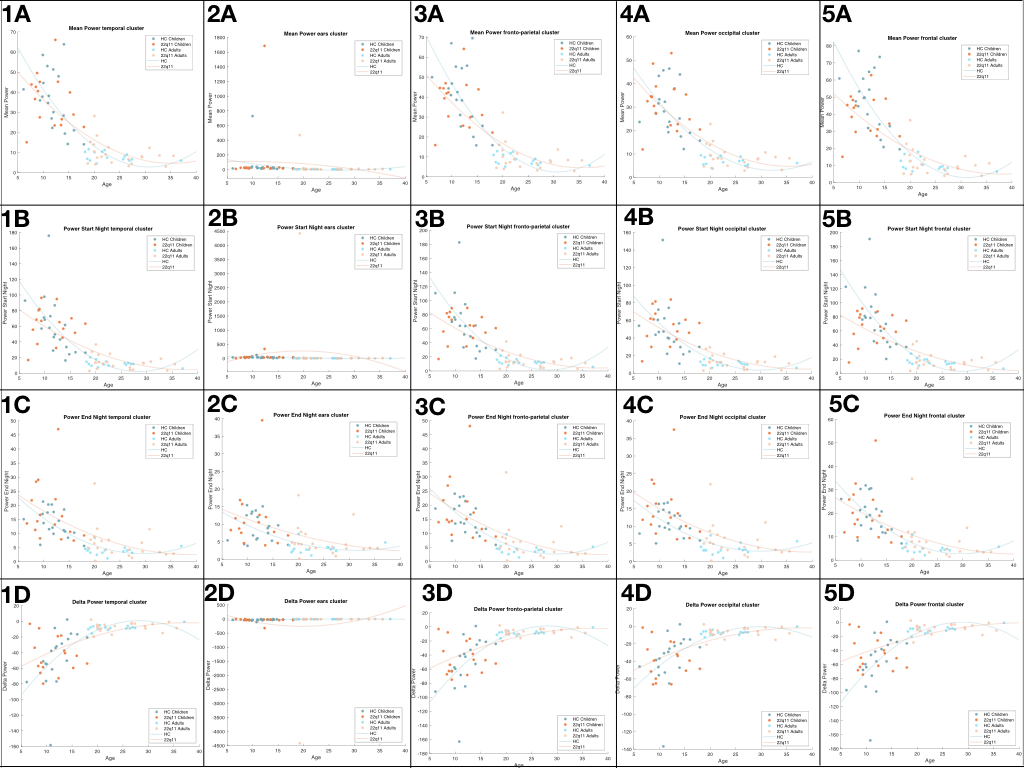

*Supplementary figure 6: Slow-waves analysis. Scatter plot of SW variable by age.* ***Panel 1:*** *Temporal cluster.* ***Panel 2:*** *Cluster around the ears.* ***Panel 3:*** *Fronto-parietal cluster.* ***Panel 4:*** *Occipital cluster.* ***Panel 5:*** *Frontal cluster* ***Panels A****: Mean power across the night.* ***Panels B:*** *Mean power of the first 30 min of NREM sleep.* ***Panels C:*** *Mean power of the last 30 min of NREM sleep.* ***Panels D:*** *Delta power (last 30 min – first 30 min).*

|  | Cluster | **HC Children** | **22q11DS Children** | **HC Adults** | **22q11DS Adults** |
| --- | --- | --- | --- | --- | --- |
| *Mean Power*  mean (std) | **Temporal** | 35.99 (14.67) | 36.09 (12.99) | 9.5 (2.24) | 9.5 (5.93) |
|  | **Ears** | 60.43 (162.1) | 125.6 (415.95) | 29.42 (1.63) | 29.42 (101.22) |
|  | **Fronto-parietal** | 40.39 (16.04) | 37.09 (12.13) | 9.6 (2.64) | 9.6 (6.61) |
|  | **Occipital** | 28.8 (10.65) | 30.9 (10.77) | 8.79 (1.81) | 8.79 (4.77) |
|  | **Frontal** | 47.74 (17.31) | 39.13 (12.03) | 11.37 (3.68) | 11.37 (7.65) |
| *Start Night Power*  mean (std) | **Temporal** | 59.8 (37.05) | 61.44 (21.81) | 13.63 (3.67) | 13.63 (8.66) |
|  | **Ears** | 39.78 (24.64) | 56.19 (75.94) | 220.1 (2.9) | 220.1 (962.27) |
|  | **Fronto-parietal** | 66.98 (39.15) | 62.84 (21.24) | 13.78 (3.96) | 13.78 (9.37) |
|  | **Occipital** | 48.58 (30.48) | 53.47 (19.87) | 12.76 (3.17) | 12.75 (7.5) |
|  | **Frontal** | 76.8 (40.15) | 65.16 (21.59) | 16.26 (5) | 16.26 (11.01) |
| *End Night Power*  mean (std) | **Temporal** | 13.73 (4.79) | 17.3 (10.47) | 6.31 (1.62) | 6.31 (5.8) |
|  | **Ears** | 9.08 (3.99) | 11.37 (8.39) | 5.1 (1.43) | 5.1 (4.12) |
|  | **Fronto-parietal** | 15.32 (5.61) | 17.69 (10.41) | 6.44 (1.82) | 6.44 (6.57) |
|  | **Occipital** | 11.19 (3.6) | 14.59 (7.89) | 5.8 (1.58) | 5.8 (4.66) |
|  | **Frontal** | 19.36 (7.83) | 19.61 (11.16) | 7.46 (2.04) | 7.46 (7.21) |
| *Delta Power*  mean (std) | **Temporal** | -46.07 (36.02) | -44.14 (22.73) | -7.32 (3.26) | -7.32 (5.38) |
|  | **Ears** | -30.7 (23.42) | -44.82 (75.63) | -215.01 (2.45) | -215.01 (962.42) |
|  | **Fronto-parietal** | -51.65 (37.67) | -45.15 (22.87) | -7.33 (3.45) | -7.33 (5.27) |
|  | **Occipital** | -37.39 (29.84) | -38.89 (20.89) | -6.95 (2.81) | -6.96 (5.45) |
|  | **Frontal** | -57.43 (38.21) | -45.55 (23.12) | -8.81 (4.49) | -8.81 (6.9) |

Supplementary table 3: *Slow-waves variables across the night by group and age, electrodes were averaged by clusters. Mean (std)*

1.	Alsubie, H.S. and A.S. BaHammam, *Obstructive Sleep Apnoea: Children are not little Adults.* Paediatric Respiratory Reviews, 2017. **21**: p. 72-79.
